# How to tie dangerous surgical knots – easily. Can we avoid this?

**DOI:** 10.1101/2021.04.12.21255295

**Authors:** E H Drabble, S Spanopoulou, E. Sioka, E Politaki, I K Paraskeva, E Palla, L Stockley, D Zacharoulis

**Affiliations:** University of Thessaly, Greece; Clinical lead BSS course RCS England, Consultant Surgeon, University of Plymouth Hospitals NHS Trust; University of Plymouth, England

**Keywords:** Knot security, Suture material, Flat reef knot (FRK), Knots tied under tension (TK), Knots laid without appropriate hand crossing (NHCK)

## Abstract

**Objective:** Secure knots are essential. Previous publications have concentrated on security of different knot types, but could individual technique be important?

Determine whether the technique of formation of each layer of a surgical knot is important to the security of the knot formed.

**Design study:** Prospective analysis of technique on knot security

**Materials and methods:** Senior and resident surgeons, and medical students, tied knots with three techniques, using four study materials, 2/0 polyglactin 910 (vicryl), 3/0 polydioxanone (PDS), 4/0 poliglecaprone 25 (monocryl) and 1 nylon (Ethilon); a standard flat reef knot (FRK), knots tied under tension (TK), and knots laid without appropriate hand crossing (NHCK). Each knot technique was performed reproducibly, and security determined by distraction with increasing force, till each material broke, or the knot separated completely.

**Results:** 20% of flat reef knots (FRK) tied with all suture materials slipped; all knots tied with the other two techniques, with all materials, slipped, TK (100%) and NHCK (100%). The quantitative degree of slip, was significantly less for FRK (mean 6.3% 95%CI 2.2-10.4%) than for TK (mean 312% 95%CI 280.0-344.0%) and NHCK (mean 113.0% 95%CI 94.3-131.0%).

The mean lengths of suture in loops held within knots, tied under tension (TK mean 17.0mm 95%CI 16.3-17.7mm), and tied without appropriate hand crossing (NHCK mean 16.3mm 95%CI 15.9-16.7mm) were significantly lower than for flat reef knots (FRK mean 25.1mm 95%CI 24.2-26.0mm). The first two types of knot may have tightened more than anticipated, in comparison to flat reef knots, with potential undue tissue tension.

**Conclusion:** Meticulous technique of knot tying, is essential for secure knots, appropriate tissue tension, and the security of anastomoses and haemostasis effected.

**Strengths and limitations:** The study design was simple, with equal numbers of knots tied for each technique, and for each material.

Only a small number of participants tied knots, limiting any assessment to the effect of the knot tying technique. No inference could be drawn regarding the effect of seniority of participant.

A relatively small number of knots were tied with each of the materials limiting more detailed assessment of the effect of the suture material and size on knot security, or whether material and size had any significant influence.

## Introduction

Knot tying is an essential basic practical skill required by all surgeons, veterinary surgeons, and any clinician engaged in patient procedures, in all medical as well as surgical specialities. Secure knots that will not slip or fail, are essential for safe surgical and interventional practice, ensuring haemostasis, the integrity of anastomoses, secure and appropriate apposition of wounds, and security of interventional devices. A number of papers have investigated what type of knot could be considered to be best, even looking at the addition of surgical glue to aid security *(1-44)*. What has not been assessed, in objective detail, is the influence of the actual technique of formation of each layer of the knot on the integrity and security of a square surgical knot, rather than what type of knot.

The security of a knot tied with any material relies on the friction between layers of material applied to make the knot, and the greater the lengths of both sections of suture brought firmly together to entwine and hold against each other, the greater the friction and security of the knot (1,*45,46,)*.

The advent of more modern suture materials, and more monofilament sutures, has led us to apply more layers of material, or more “throws”, to create secure knots as these materials are considered to be more slippery than older materials, and the current recommendation is that we should apply at least six layers of material when securing knots with a monofilament material, such as polypropylene or nylon*(2,3,13,22,47,48,49,50,51)*. However, the number of “throws” or layers of suture laid in each knot could be irrelevant if the technique with which the knot is formed is inadequate. *(1,14,37,38,39)*

This initial study assessed the impact of technique of knot formation on the integrity and security of standard surgical reef knots, tied using four commonly used suture materials of varying thickness, or strength, and performed by three grades of surgeon, a senior consultant, a surgical trainee who had previously been taught on the Intercollegiate Basic Surgical Skills course (*52*) and three medical students. The intention was to determine the influence of technique on the integrity of surgical reef knots, the most commonly used form of knot in surgical procedures; size and strength of the suture material, and experience of the operating surgeon were considered to be less likely to be important. (*20,27,29,31,34,41,48,49,50,53,54*)

## Materials and Methods

Three techniques of tying a surgical reef knot were determined and designed so that each participating surgeon would tie each of the three knots in a reproducible manner common to all participating surgeon groups. Each knot was tied using a needle holder, with instrument tying techniques to facilitate reproducibility between all three surgeon groups.

The first technique was creation of a flat reef knot (FRK) with each layer of the knot, or “throw”, formed with equal and opposite movements of the hands and needle holder, so that each hand crossed each other at an angle of 180 degrees, placing each layer of the knot at precisely the same level, or plane, as the knot itself, ensuring that equal amounts of each end of the suture material used were placed and intertwined in a flat horizontal layer (Figure 1).

**Figure 1.**
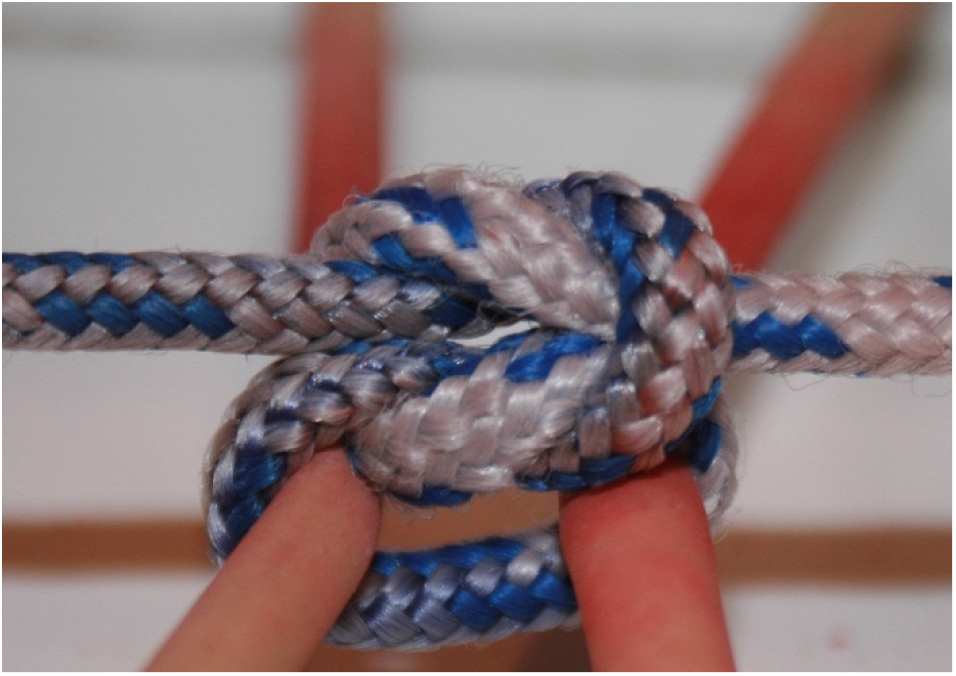

The second technique was designed to mimic a method where some surgeons maintain tension on a knot to try and prevent it potentially loosening during tying, such as that employed by some when tying a knot at depth (TK). This is usually performed by keeping one end of the suture material stiff to maintain tension on the knot, whilst forming the knot predominantly with the other end. This was performed in this study by keeping one end of the suture tense in a vertical plane, but moving the other end of the suture material producing equal and opposite movements across the knot so that each hand movement was at an angle of 180 degrees to the other, but only with one hand rather than both (Figure 2). All layers were placed in the same horizontal plane.

**Figure 2.**
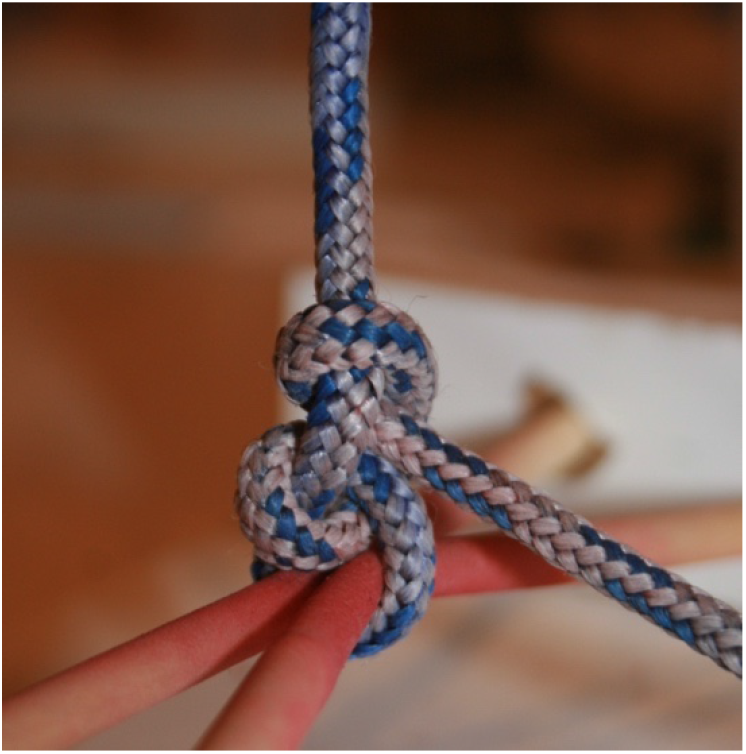

The third technique was designed to mimic a type of mistake where the operating surgeons forgets, or neglects, to remember to cross their hands with each layer of the knot so they neglect to produce equal and opposite hand movements with each layer of the knot (NHCK). Each surgeon would diligently form each layer of the knot as if they intended to perform equal and opposite movements of the hand, alternating formation of layers that should be laid in a downward direction with those that should be laid in the opposite upward direction, but each layer was completed with one hand always moving in a downward direction towards the surgeon (Figure 3) (Figure 4). All layers of the knot were placed in one horizontal plane perpendicular to the knot.

**Figure 3.**
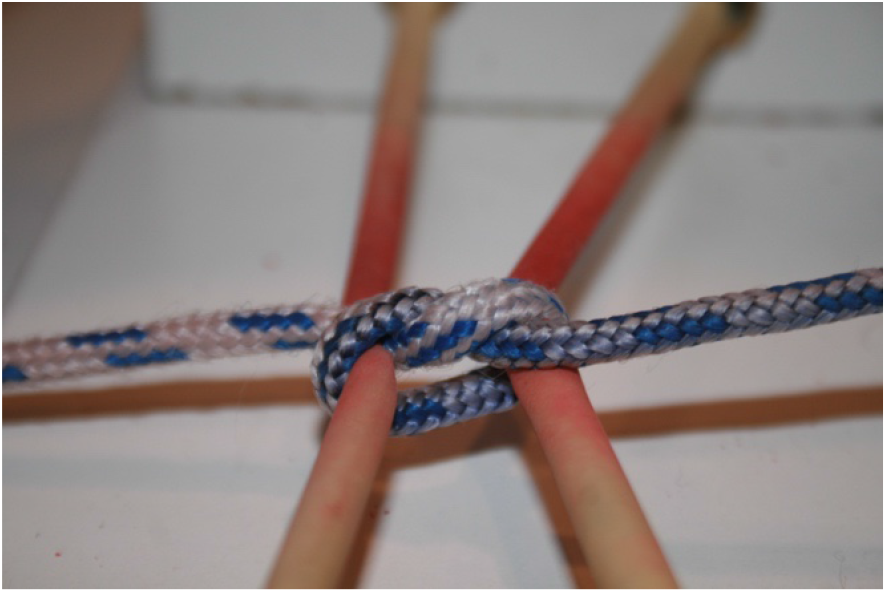

**Figure 4.**
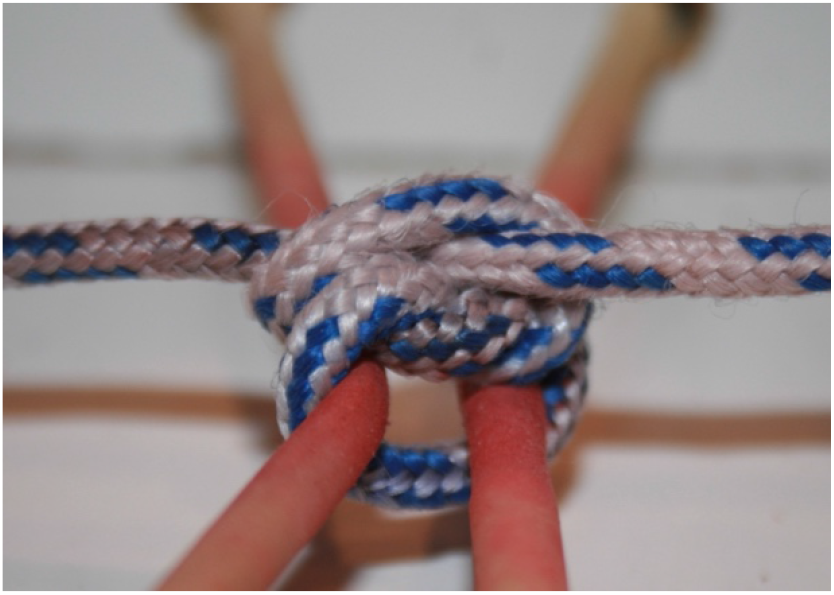

A senior consultant surgeon, a junior surgeon, and a group of three medical students, each tied ten knots of each of the three techniques, using four suture materials of different calibres, three monofilament, 4/0 poliglecaprone 25 (monocryl), 3/0 polydioxanone (PDS) and No.1 nylon (Ethilon), and one braided suture, 2/0 polyglactin 910 (vicryl), provided by Ethicon Greece. All performed these knots at one centre, on one study day. Each knot was tied across an apparatus designed to test its strength and integrity, analogous to that used in other studies on strength of knots (*23,29,55,56*). This consisted of a chain attached to a fixed clamp which was then tied to a spring loaded weight measuring device that allowed incremental increases in the weight force applied to the knot and measurement of it (Figure 5).

**Figure 5.**
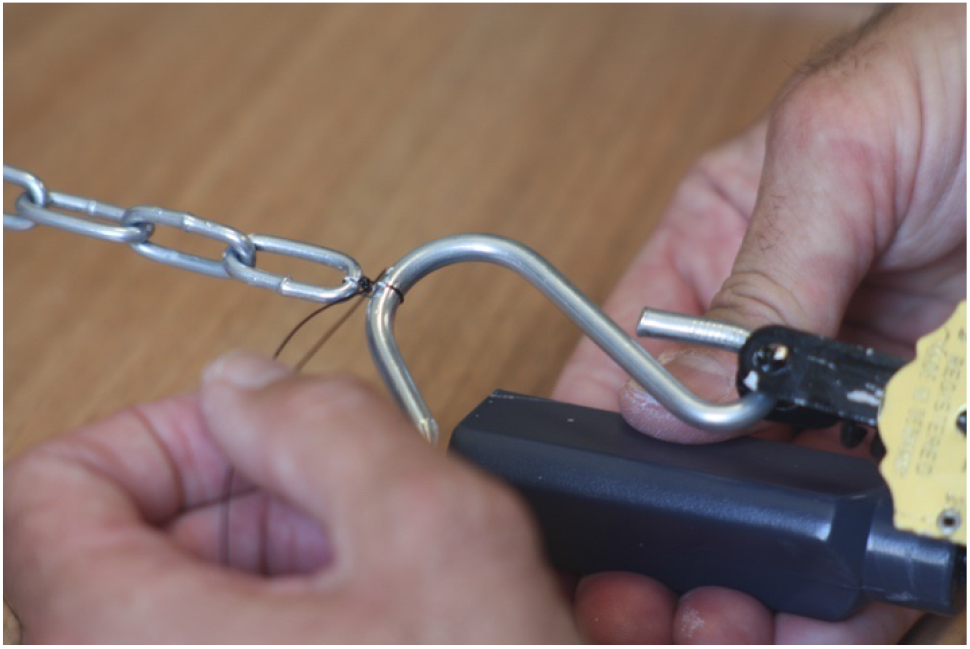
Equipment for measuring force applied to knot.

Once the knot was created, each end of the knot was marked with an indelible marker, rapid drying Tippex marker for 2/0 polyglactin 910 (vicryl), 3/0 polydioxanone (PDS) and No. 1 nylon (Ethilon) sutures, and blue indelible dye for 4/0 poliglecaprone 25 (monocryl) sutures. The pressure applied to each knot was increased incrementally till one of three final events occurred; the knot breaks completely with no evidence of slippage of the knot prior to rupture, slippage of the knot and then rupture of the suture, or complete failure of the knot as it slips and completely unravels. Each outcome was recorded, and each suture was photographed following its final outcome. The force at which each knot broke or slipped was measured. The degree by which a knot slipped when tested, was determined by measuring the amount of suture material that appeared, between the indelible markers applied to each end of the suture as it met the knot prior to testing, and the knot itself following application of force (Figure 6) (Figure 7). The amount of material that appeared at either end of the knot following application of sufficient force to break it, or cause it unravel, was measured using digital calipers, compared to the length of suture material included in the loop held by the original knot, and expressed as a percentage proportion of the length of suture included in that loop.

**Figure 6.**
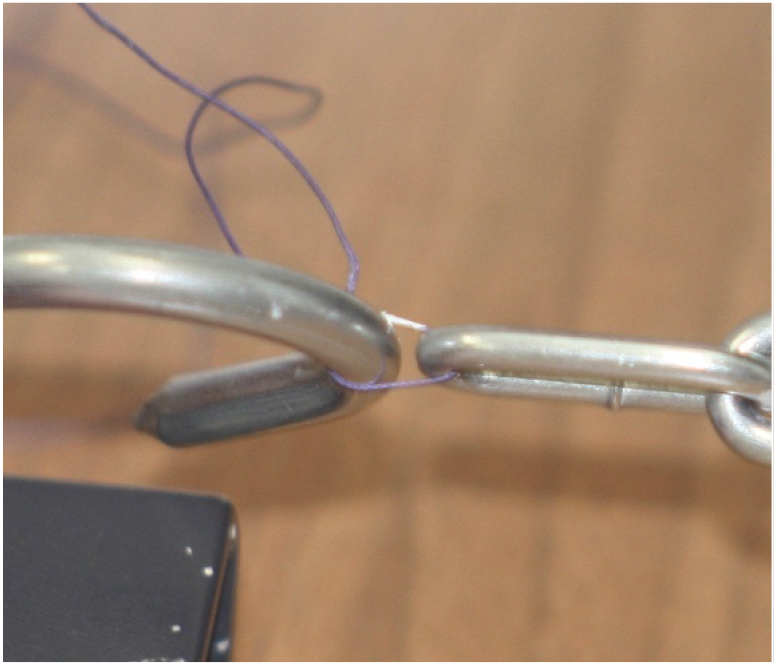
demonstrating length of material incorporated in loop held by the knot, prior to breakage on testing, and minimal slippage as demonstrated by suture material beyond white marker on left side (figure 7) post rupture suture

**Figure 7.**
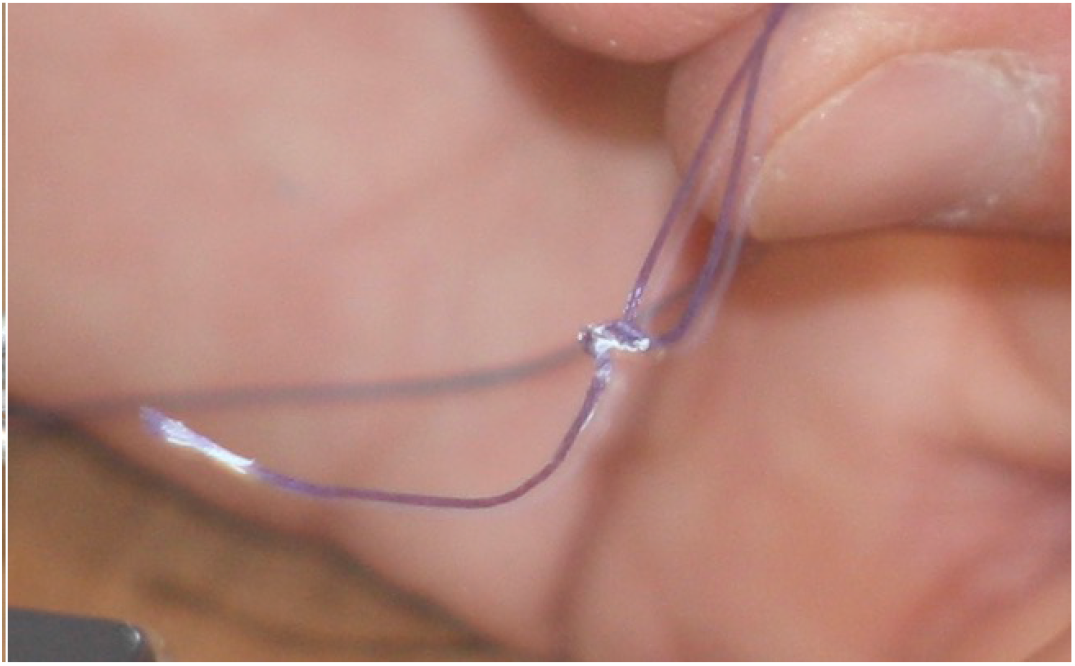
demonstrating length of material incorporated in loop held by the knot, prior to breakage on testing, and minimal slippage as demonstrated by suture material beyond white marker on left side (figure 7) post rupture suture

The force required to break each knot, or cause it to slip and unravel, was measured using the spring loaded weight measuring device and expressed in kilograms force.

The results obtained were tested for statistical significance by determining their means and 95% confidence intervals (95% CI).

## Results

Equal numbers of each type of knot technique were tied (120) **(Table 1)**, equal numbers of knots by each surgeon group (120) **(Table 1)**, and equal amounts of each suture material were used with each technique (90). **(Table 2)**. Each surgeon group tied 10 knots of each of the three knot types, with each of the four suture materials, producing a total of 360 knots for testing and assessment.

**Table 1:**
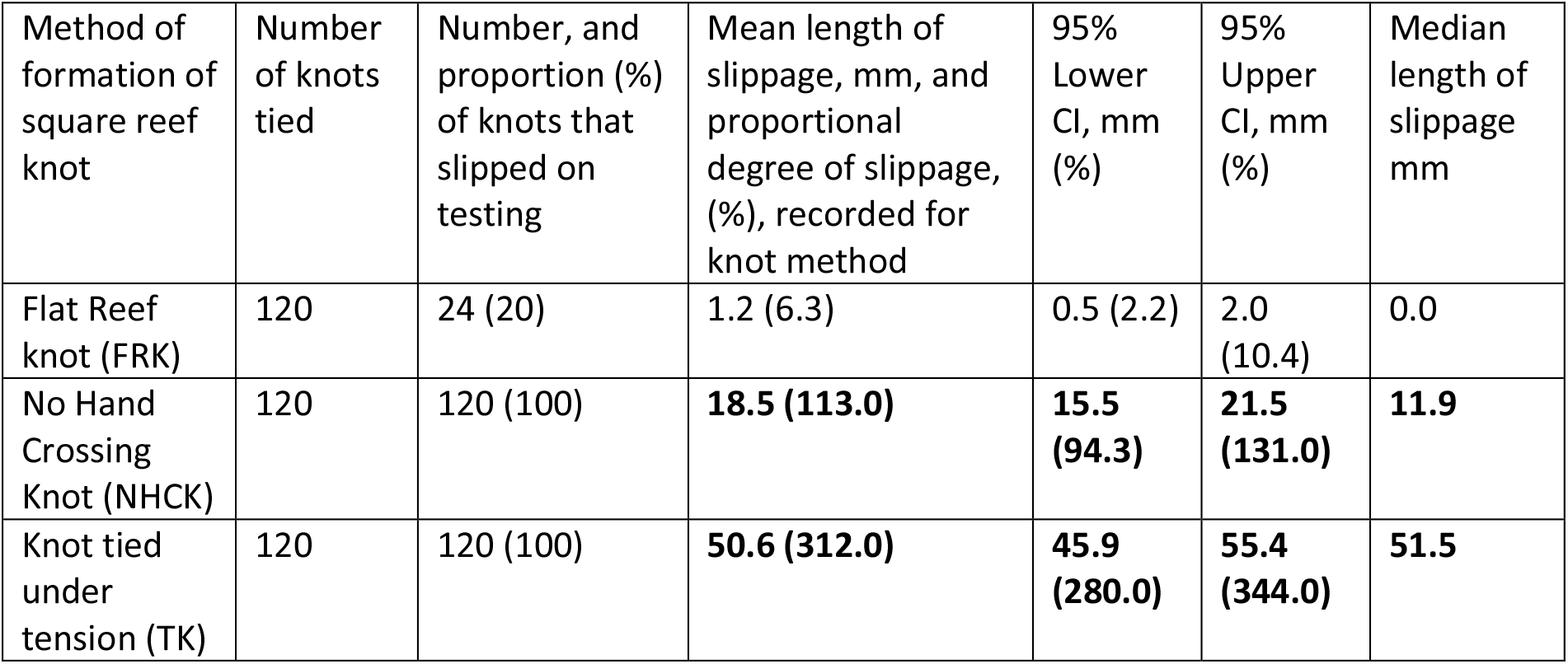
Number of knots tied with each method, and proportion of knots tied with each method, that slipped on testing, mean length of slippage mm, and proportional increase in amount of suture material held within knot post slippage, for each method, and 95% confidence intervals

| Method of formation of square reef knot | Number of knots tied | Number, and proportion (%) of knots that slipped on testing | Mean length of slippage, mm, and proportional degree of slippage, (%), recorded for knot method | 95% Lower CI, mm (%) | 95% Upper CI, mm (%) | Median length of slippage mm |
| --- | --- | --- | --- | --- | --- | --- |
| Flat Reef knot (FRK) | 120 | 24 (20) | 1.2 (6.3) | 0.5 (2.2) | 2.0 (10.4) | 0.0 |
| No Hand Crossing Knot (NHCK) | 120 | 120 (100) | **18.5 (113.0)** | **15.5 (94.3)** | **21.5 (131.0)** | **11.9** |
| Knot tied under tension (TK) | 120 | 120 (100) | **50.6 (312.0)** | **45.9 (280.0)** | **55.4 (344.0)** | **51.5** |

**Table 2:** Number of knots tied with each suture material, proportion of each that slipped on testing, mean degree of slippage in mm and proportional increase in amount of suture material held within each knot after slippage (%), for each suture material and 95% CI

| Suture material | Number of knots tied with suture material | Number and proportion of knots that slipped n (%) | Mean length of slippage of knots tied with each suture material mm, and proportional increase in length of suture material held within knot post slip (%) | Lower 95% CI of mean length and mean proportion of slippage mm (%) | Upper 95% CI of mean length and mean proportion of slippage mm (%) | Median slippage mm |
| --- | --- | --- | --- | --- | --- | --- |
| 2/0 polyglactin (Vicryl) | 90 | 71 (78.9%) | 24.2 (136.0) | 18.6 (104.0) | 29.8 (167.0) | 14.0 |
| 4/0 poliglecaprone 25 (Monocryl) | 90 | 60 (66.7%) | 15.5 (108.0) | 10.9 (73.8) | 20.2 (142.0) | 4.0 |
| 3/0 polydioxanone (PDS) | 90 | 63 (70.0%) | 19.1 (119.0) | 13.6 (83.0) | 24.5 (154.0) | 8.6 |
| 1 nylon (Ethilon) | 90 | 70 (77.8%) | 34.9 (213.0) | **28.6 (173.0)** | **41.2 (252.0)** | **27.7** |

### Knot slippage

20% of knots tied with all suture materials, by all surgeon groups, using a flat reef knot technique (FRK) with both hands crossing at 180 degrees to each other, and all layers of the knot laid in the same plane as the knot, subsequently slipped to some degree on testing. 100% of knots tied with one hand maintaining tension on the knot (TK), and 100% of those tied with one hand always moving in a downward direction (NHCK), slipped. In addition, the mean degree of slippage, as measured as a proportional increase in the amount of material that appeared between the indelible markers and the knot itself, or as the mean length of extra material measured in mm, following formation of a flat reef knot (FRK) **(6.3% 95% CI 2.2 – 10.4%) (1.2 mm 95% CI 0.5 – 2.0 mm)**, was significantly less than the mean degrees of slippage of knots formed with the other two techniques, with one hand holding the knot under tension (TK) **(312.0% 95% CI 280.3 – 343.7%) (50.6mm 95% CI 45.9 – 55.4 mm)**, and with one hand always moving in a one direction, rather than alternately crossing the knot (NHCK) **(112.8% 95% CI 94.3 – 131.4%, 18.5 mm 95% CI 15.5 – 21.5 mm), (Table 2, plot diagram 1)**

**Plot diagram 1:**
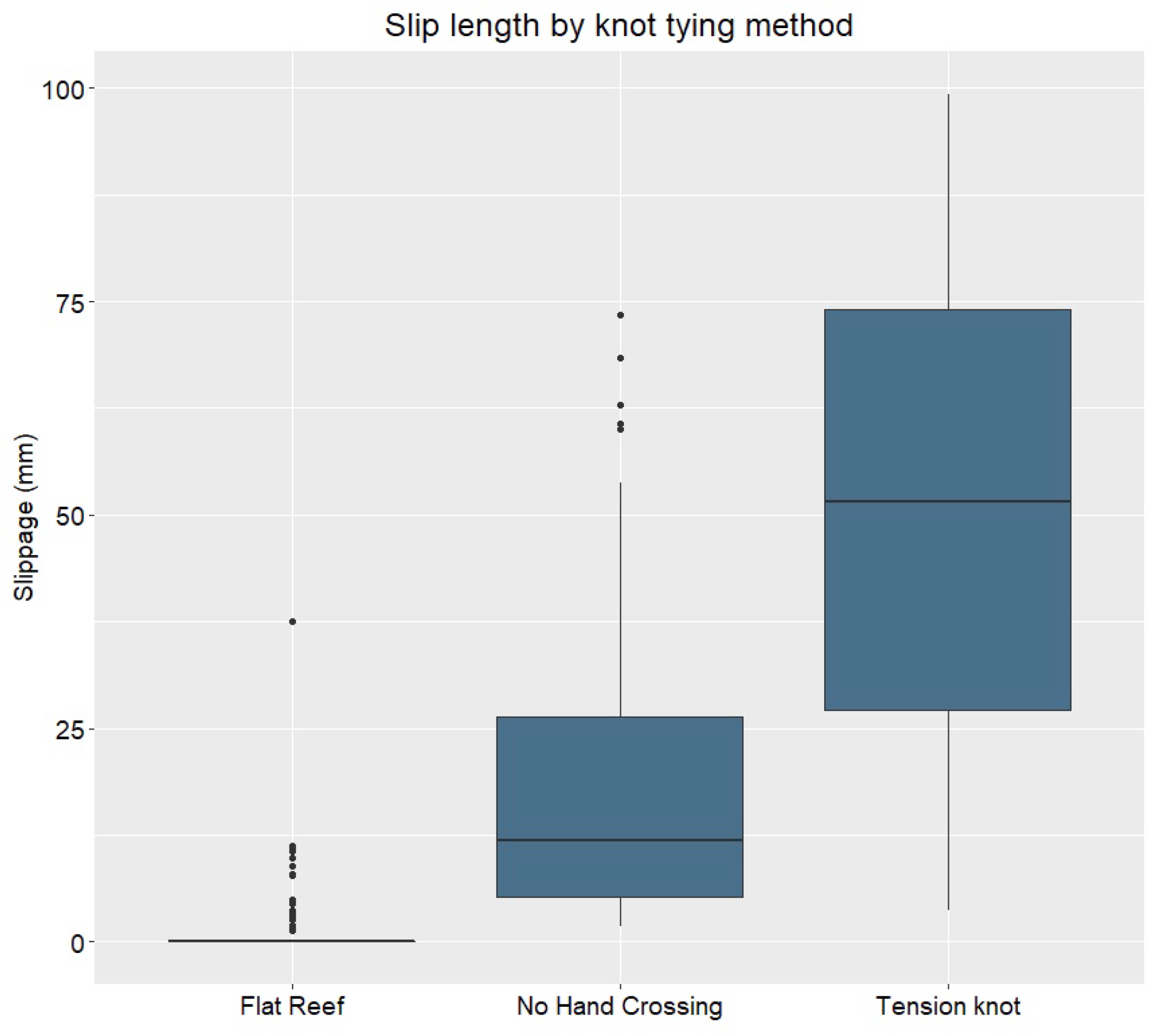
Box plot displaying the slippage length, and median length and interquartile range of slippage, in mm by knot tying method

Of the 90 knots tied with each material, the following numbers and proportions of knots slipped on testing: 2/0 polyglactin (Vicryl) 71 (78.9%), 4/0 poliglecaprone 25 (Monocryl) 60 (66.7%), 3/0 polydioxanone (PDS) 63 (70.0%), and 1 nylon (Ethilon) 70 (77.8%) **(Table 2)**. Similar proportions of the larger suture materials, 2/0 polyglactin (Vicryl) and 1 nylon (Ethilon), slipped on testing. The proportions of knots tied with the smaller diameter suture materials, 4/0 poliglecaprone 25 (Monocryl) and 3/0 polydioxanone (PDS) appeared to be smaller, but the differences were not significant.

The amounts of slippage of knots tied with the four different materials, as measured by a proportional increase in material held within the knot, did show that those tied with 1 nylon (Ethilon) slipped by a greater length, if they did slip **(mean 213.0%, 95% CI 173.0 – 252.0%)**, than those tied with the smaller diameter suture materials, 4/0 poliglecaprone 25 (Monocryl) **(mean 108.0%, 95% CI 73.8 – 142.0%)** and 3/0 polydioxanone (PDS) **(mean 119.0, 95% CI 83.0 – 154.0%)**. No solid conclusion could be drawn regarding the difference in slippage length with knots tied with 2/0 polyglactin (Vicryl). (**Table 2, plot diagram 2)**.

**Plot diagram 2:**
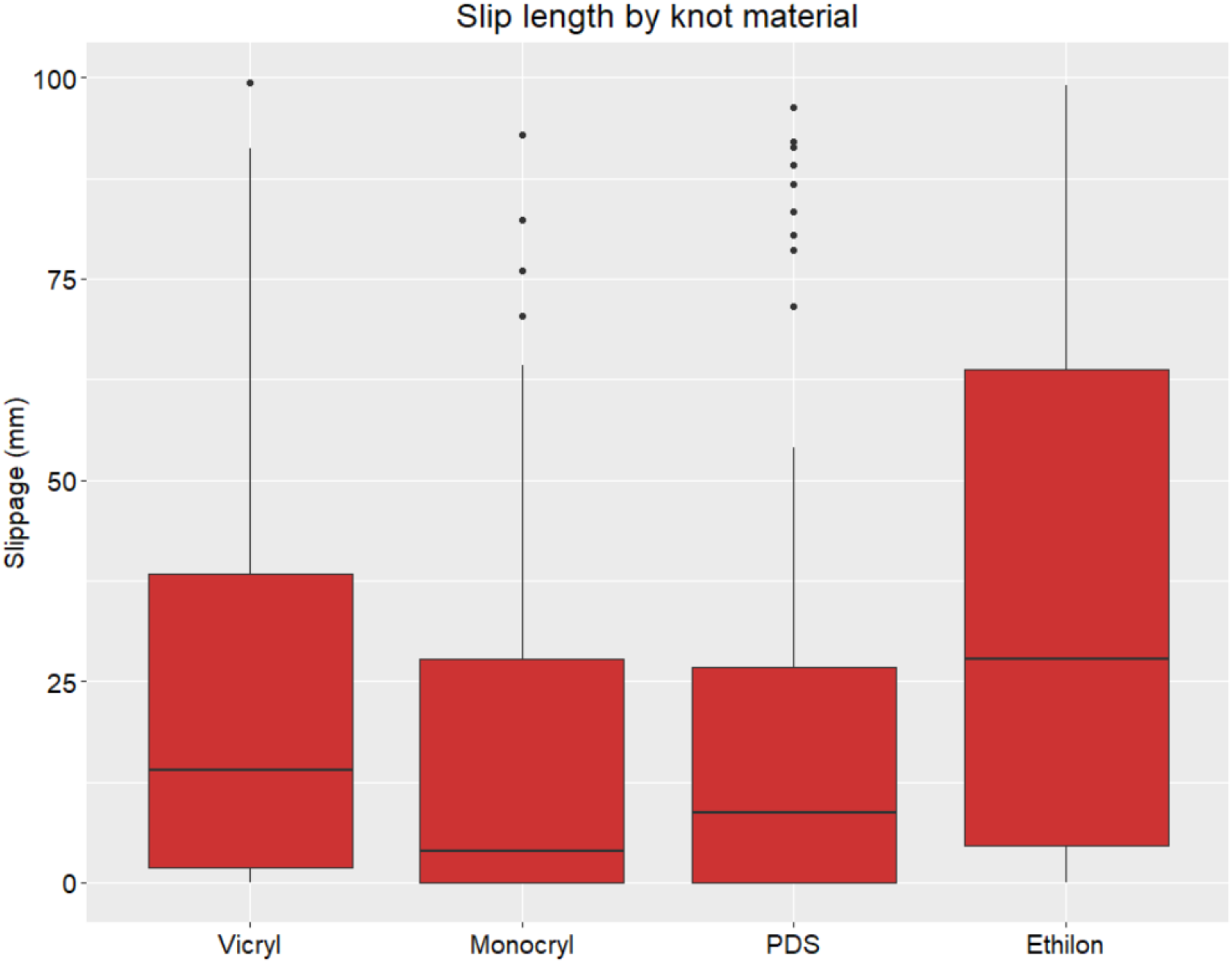
Box plot displaying lengths of slippage, and median slippage and interquartile range of slippage, for knots tied with each suture material

We assessed whether the type of suture material would have an effect on the efficacy of each knot type, determining the mean slippage length within each method for each material type.

No knots tied with 4/0 poliglecaprone 25 (Monocryl) using the flat reef knot technique slipped to any appreciable extent. We observed no significant difference between the lengths of slippage of flat reef knots tied with the other three suture materials. Knots tied with 1 nylon (Ethilon) using the technique of not crossing your hands appropriately, and those tied whilst keeping the knot under tension, did appear to slip more than knots tied with the same techniques, using the other three suture materials. **(Table 3, plot diagram 3)**

**Table 3:** Number of knots tied with each suture material, using each technique, and length of slippage mm

| Method | Material | Number of knots tied with technique and suture material | Number of knots that slipped | Mean (mm) | Lower CI | Upper CI |
| --- | --- | --- | --- | --- | --- | --- |
| Flat Reef Knot (FRK) | 2/0 polyglactin (Vicryl) | 30 | 11 (36.7%) | 1.4 | 0.4 | 2.4 |
| Flat Reef Knot (FRK) | 4/0 poliglecaprone 25 (Monocryl) | 30 | 0 (0%) | **0.0** | **0.0** | **0.0** |
| Flat Reef Knot (FRK) | 3/0 polydioxanone (PDS) | 30 | 3 (10%) | 0.3 | -0.1 | 0.6 |
| Flat Reef Knot (FRK) | 1 nylon (Ethilon) | 30 | 10 (33.3%) | 3.3 | 0.5 | 6.0 |
| No Hand Crossing Knot (NHCK) | 2/0 polyglactin (Vicryl) | 30 | 30 (100%) | 19.9 | 15.1 | 24.8 |
| No Hand Crossing Knot (NHCK) | 4/0 poliglecaprone 25 (Monocryl) | 30 | 30 (100%) | 7.1 | 2.5 | 11.7 |
| No Hand Crossing Knot (NHCK) | 3/0 polydioxanone (PDS) | 30 | 30 (100%) | 13.6 | 9.0 | 18.3 |
| No Hand Crossing Knot (NHCK) | 1 nylon (Ethilon) | 30 | 30 (100%) | **33.2** | **26.9** | **39.6** |
| Knot tied under tension (TK) | 2/0 polyglactin (Vicryl) | 30 | 30 (100%) | 51.4 | 41.5 | 61.2 |
| Knot tied under tension (TK) | Monocryl4/0 poliglecaprone 25 (Monocryl) | 30 | 30 (100%) | 39.6 | 31.7 | 47.5 |
| Knot tied under tension (TK) | 3/0 polydioxanone (PDS) | 30 | 30 (100%) | 43.4 | 32.1 | 54.6 |
| Knot tied under tension (TK) | 1 nylon (Ethilon) | 30 | 30 (100%) | **68.3** | **62.2** | **74.4** |

We believe this is likely to be due to this large monofilament suture being stronger and less likely to break under tension than smaller diameter suture materials. Knots tied with this material that then slip are less likely to break than those tied with the other materials. **(Table 5)**

**Plot diagram 3:**
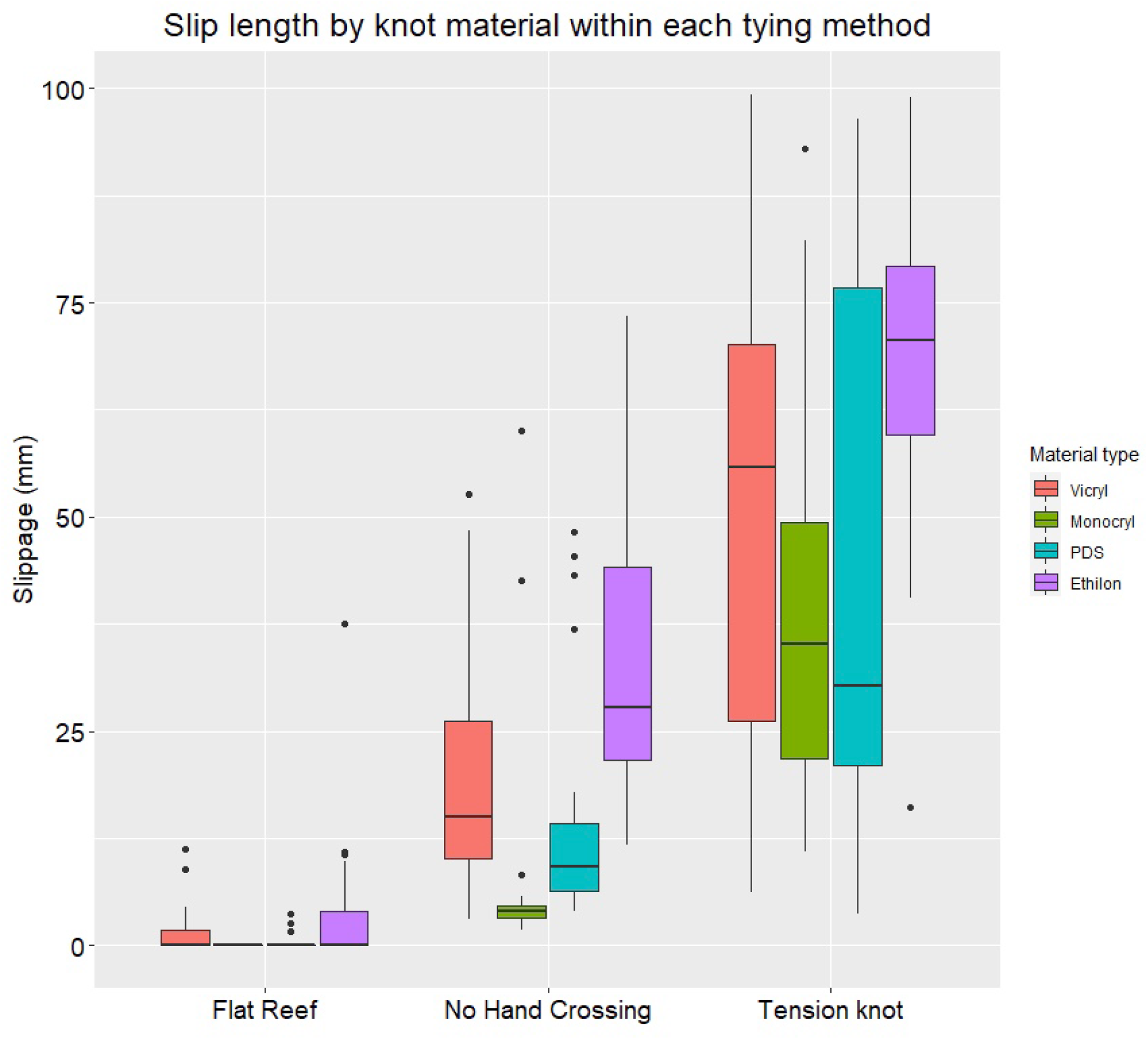
Box plot of slip lengths of knots tied with each of the three techniques, using the four suture materials

### Knot breakage

Of 360 knots tied, 284 broke on testing, with or without some degree of slippage, 76 slipped completely without breaking.

The proportion of knots that broke completely on testing, whether following some degree of slippage prior to breaking or not slipping at all, was significantly greater for those tied with a flat reef knot technique (FRK) **(0.99, 95% CI 0.98 – 1.00)**, than those tied with one hand holding the knot under tension (TK) **(0.56, 95% CI 0.47 – 0.65)**, and with one hand always moving in a downward direction (NHCK) **(0.82, 95% CI 0.75 – 0.89)** One knot of 120 tied with a flat reef knot technique (FRK) did slip completely, but this compares favourably with 53 knots tied with one hand holding the knot under tension that slipped completely (TK), and 22 of those tied with one hand moving in a downward direction for all layers of the knot (NHCK). **(Table 4)**

**Table 4:** Number of knots tied using each knot technique that slipped and broke and slipped completely, and number and proportion that broke on testing

| Method of formation of square reef knot | Total number of knots formed that slipped | Number of knots that slipped and then broke | Number of knots that slipped completely without any suture breakage | Proportion of knots formed that broke and did not slip completely | 95% lower CI of proportion | 95% upper CI of proportion |
| --- | --- | --- | --- | --- | --- | --- |
| Flat Reef knot (FRK) | 24 | 23 | 1 | 0.99 | 0.98 | 1.00 |
| No Hand Crossing Knot (NCHK) | 120 | 98 | 22 | **0.82** | **0.75** | **0.89** |
| Knot tied under tension (TK) | 120 | 67 | 53 | **0.56** | **0.47** | **0.65** |

On assessing what proportions of knots tied using each of the four suture materials employed in this study, virtually all knots tied with 4/0 poliglecaprone 25 (Monocryl) broke on testing **(87, 96.7%)**. Fewer of those tied with 2/0 polyglactin (Vicryl) **(74, 82.2%)** and 3/0 polydioxanone (PDS) **(76, 84.4%)** broke, and only just over half of those tied with 1 nylon (Ethilon) broke **(47, 52.2%). (Table 5)**

**Table 5:** Number and proportion of knots tied using each suture material that broke on testing

| Suture material used for knot formation | Number of knots formed | Number of knots that broke on testing | Proportion of knots formed that broke on testing % |
| --- | --- | --- | --- |
| 2/0 polyglactin (Vicryl) | 90 | 74 | 82.2 |
| 4/0 poliglecaprone 25 (Monocryl) | 90 | 87 | 96.7 |
| 3/0 polydioxanone (PDS) | 90 | 76 | 84.4 |
| 1 nylon (Ethilon) | 90 | **47** | **52.2** |

### Length of suture material included in each knot type prior to testing

The mean lengths of suture material incorporated into knots, that is the length of material in the loop tied around the hooks held by the knot was measured for all 360 knots. The average lengths of suture material included in the loop for knots tied under tension **(TK mean 17.0mm 95%CI 16.3-17.7mm)**, and those tied without the operator crossing their hands **(NHCK mean 16.3mm 95%CI 15.9-16.7mm)** were significantly lower than that for flat reef knots (**FRK mean 25.1mm 95%CI 24.2-26.0mm). (Table 6)**. This would suggest that that the first two types of knot may tighten more than anticipated, once they are initially formed, in comparison to flat reef knots, and this further tightening may potentially produce undue tissue tension, which may affect tissue viability and healing.

**Table 6:** Mean lengths of suture material incorporated into loop holding hooks in test bed, for each type of knot method

| Method of formation of square reef knot | Total number of knots formed | Mean length of suture material incorporated into loop held by knot mm | 95% lower CI of mean | 95% upper CI of mean |
| --- | --- | --- | --- | --- |
| Flat Reef knot (FRK) | 120 | 25.1 | 24.2 | 26.0 |
| No Hand Crossing Knot (NCHK) | 120 | **16.3** | **15.9** | **16.7** |
| Knot tied under tension (TK) | 120 | **17.0** | **16.3** | **17.7** |

## Discussion

The ability to tie a reliable and secure knot has always been, and remains, an essential skill for any surgeon, veterinary surgeon, or clinician engaged in any practical clinical discipline (*1*), and even with the advent of new technological aids and robotic machines to aid our surgical practice (*64,65,66*), formation of a secure knot remains an essential part of an individual surgeon’s or practitioner’s craft. In our study, the majority of knots tied with a flat reef knot technique were secure, with no slippage at all, and of those that did slip, the proportional slip was small. Knots tied with this technique, preformed carefully to ensure equal and opposite lengths of suture were entwined, could be considered to be reliable. Those tied with the other two techniques were not secure. All of the latter two types slipped, with all of the materials used, and all tied by each surgeon group.

The degree to which these knots tied with the other two techniques slipped, was remarkable, particularly those tied with one hand maintaining tension on the knot at all times **(table 1, plot diagram 1)**. Those tied with a technique that mimicked a surgeon failing to cross their hands appropriately slipped by more than 100%, those tied by a technique mimicking a surgeon maintaining tension on the knot throughout its formation, as may occur tying in a difficult location or at depth, (*1,32,43,62*) more than 300%. In contrast, the mean degree of slip of knots tied with a flat reef knot technique was markedly less, 6.3%. The small mean result may have been influenced by the large denominator, 120 knots in total, but we did observe that the majority of those flat reef knots that did slip only slipped by a small margin.

Though we observed that knots tied with the largest diameter suture material, 1 nylon (Ethilon), once they did slip, appeared to slip to a greater extent than those tied with the other materials, **(Tables 2 and 3, plot diagrams 2 and 3)**, the proportion of knots using this material that slipped was not markedly greater **(Table 2)**. This apparent difference between suture materials was probably due to the inherent greater strength of the larger suture in comparison to the others; only 52% of knots tied with this larger suture broke on testing. **(Table 5)**.

All knots tied with 4/0 poliglecaprone 25 (Monocryl), using the flat reef knot technique, held firm and broke on testing, without slippage. **(Table 3, plot diagram 3)** *(63)*. We can make no other conclusions regarding results of individual suture materials, only that the technique used markedly influenced the security and reliability of the knot formed, for all the materials used in this study. **(Tables 1, 3 and 4, plot diagrams 1 and 3)**.

Knots tied with the techniques of maintaining tension on the knot with one hand, and those tied by an operator failing to cross their hands appropriately, included less material in the loop held by the knot than those tied with a flat reef knot technique **(table 6)**. This may be due to knots tied with these techniques being more prone to slip, and these knots may have slipped more tightly again once the first throw was laid, so less material would be subsequently left within the loop secured by the knot.

If such knots can slip more tightly again, following initial formation of the knot, this may lead to undue tension being applied to a suture, and this, in turn, could lead to undue and unintended tension on the suture. This may have a detrimental effect on tissue healing, such as a bowel or ureteric anastomosis, or wound closure. Insecure knots may cause harm not only from loosening and slipping post formation, but also from squashing and crushing tissue inadvertently during their initial formation.

The salient result was the marked difference in the integrity of knots tied with a flat reef knot technique in comparison to those using the other two techniques, and such was the degree of slippage that we should probably consider knots tied with these two other methods dangerous.

Why did these dramatic knot failures occur in our study? Was this a failure of the design of the study, are our results clinically relevant; are we using the wrong type of knot; are modern suture materials too slippery for secure knot formation; and if we accept there is a potential widespread problem with the security of surgical knots in surgical practice in general, can we overcome the problem, or rely on technology to find other ways of securing haemostasis, anastomoses and closure of wounds, other than tying a knot on a suture? (*55,64,65,66*)

We would suggest that secure and reliable surgical knots can be consistently made with the simple reef knot with modern materials, provided we employ a meticulous technique, for each and every layer, or throw, of each knot. (*41,49*)

The simple reef knot has been the most commonly used, and taught, surgical knot. The “Hercules knot”, or square knot, has been recorded to have been used in surgical practice in Greece in the first century AD, and the square knot, or reef knot has probably been used in general for 2000 years (*67,68,69*,) The reef knot, tied appropriately, was used to hold heavy, large wet sails in place, even in the worst of stormy weather, as recommended in sailing texts from the 18^th^ century, and described in instruction books for sailors in the 18^th^ and 19^th^ centuries *(70,71,72)*. If such a simple knot can control such a heavy burden in these difficult circumstances, and has been used so extensively, and for so long in surgical practice (*1,67*), why did we observe such a problem with those tied with two techniques in this study, and failure rates of 24-80% reported for reef knots tied by experienced surgeons and in studies on teaching knots to students and junior surgeons (*1,56,73,74*). The answer may lie in how reef knots are untied *(75)*, and in the nature of modern surgical suture materials.

Sailors can untie, or “unreef” a knot by pulling on one strand of the knot, this will change the configuration of the knot so that there will be unequal amounts of the two strands of rope in the knot, and one strand can adopt a straight configuration reducing its frictional surface in contact with the other strand, so that it can be undone (figures 8,9 and 10).(*45,75*)

**Figure 8.**
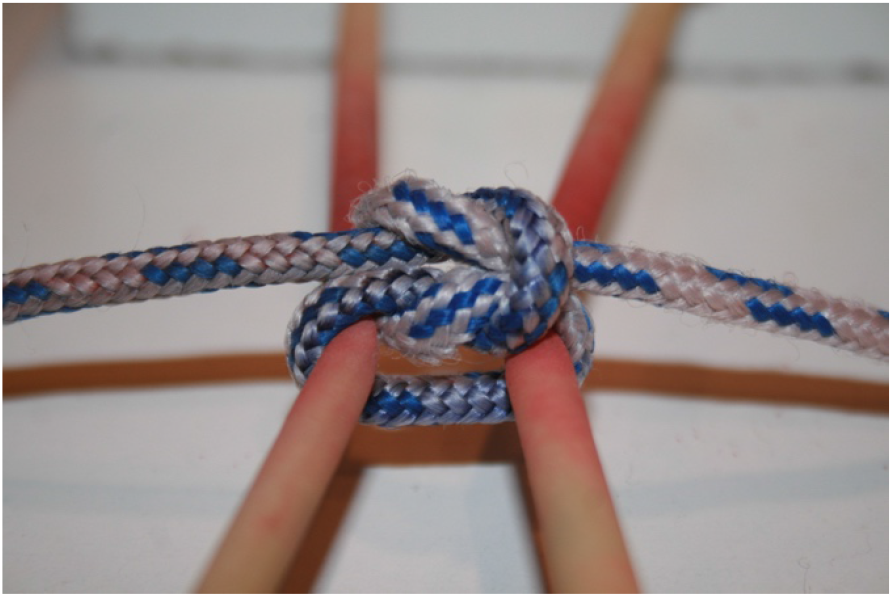

**Figure 9.**
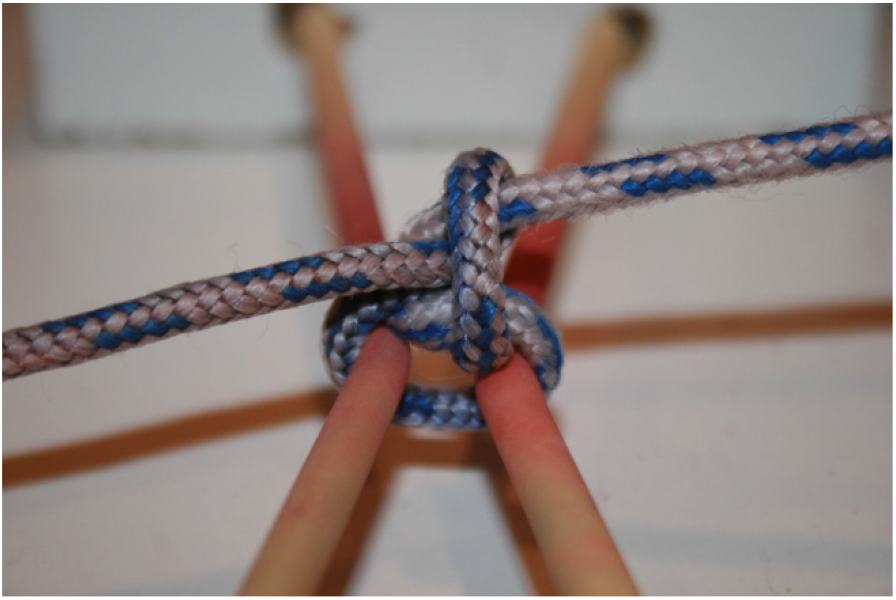

**Figure 10.**
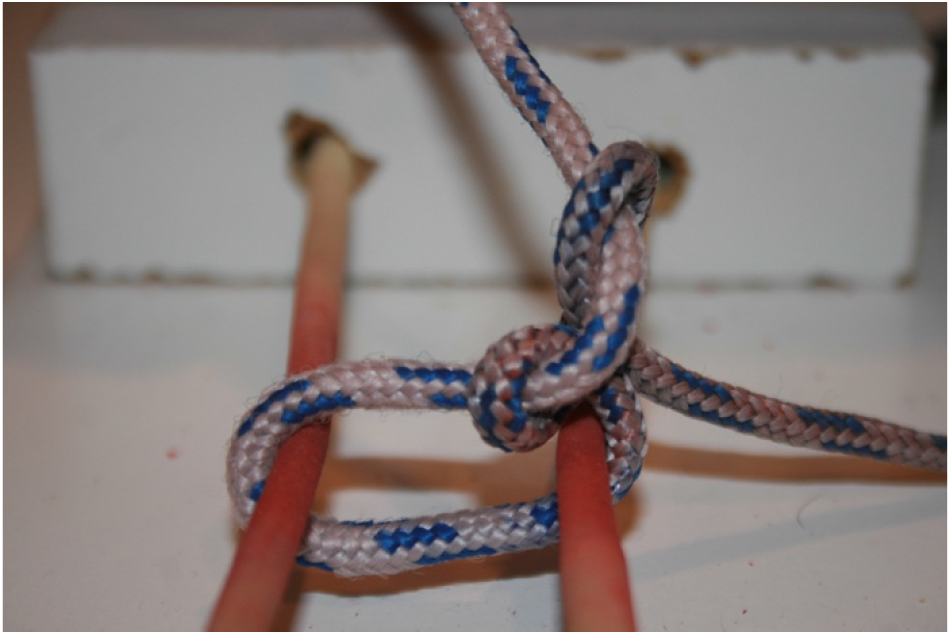

The two knot techniques that produced a failure rate of 100% would have laid unequal amounts of the two suture strands within the knot, reducing the friction between the two strands (figures 2) (Figure 4).

A knot relies on friction between the strands of material placed in mutual apposition within the knot (*1,45,46,76*), and this in turn will rely on the natural friction, or lack of slipperiness, of the suture material, and how well the two strands are laid together to produce as much mutual contact between them. The flat reef knot technique would potentially lead to as much contact as possible between the two strands, provided care is taken to ensure equal amounts of material are placed in the knot (figures 1 & 7). What we did observe, during the course of the study, and on observation of videos of the knots we formed, was that even if participants took as much care as possible in forming the flat reef knot, small twists in the material could lead to a less than perfect apposition of the suture strands at the final laying down of the knot, leading to unequal strand lengths in the final knot. This may make these particular knots less resistant to slipping on testing, and could explain why a proportion of knots tied with our flat reef knot technique slipped, and why some knots tied by senior surgeons in other studies failed.(*41,43,53*)

Knots rely on friction between the strands of material laid in the knot, and the material must have a level of surface friction sufficient to allow knots to hold (*1,46*). Though more modern suture materials would appear to more slippery, and pass through tissue less traumatically than older and traditional materials, even monofilament nylon has sufficient surface friction to allow a simple reef knot to hold.(*46*)

The study design was simple, relying on a winch applied to a weighing spring, to produce an incremental force to distract two metal hooks tied together with the four, dry, suture materials we tested. Other studies assessing different types of knots, rather than technique of knot formation, have validated the use of such simple techniques and equipment.(*2-8, 10,23,29,36,56*) We would suggest that our conclusions on knot security are clinically valid; poor technique can lead to dramatically poor results.

What should we do in clinical practice? This study, and other studies that have compared different types of knots rather than the influence of technique on knot integrity(*1-44*), have demonstrated that a proportion of knots can slip. This is a small study, and should probably be viewed as a preliminary study, but the results are clear. Do we abandon using sutures and look at other methods relying on technology, such as stapling devices for all anastomoses, haemostatic devices for haemostasis and vessel control, and staple all wounds(*55,64,65,66*), or do we accept that technique and craft are essential for a successful outcome when forming knots (*41,53*)? Relying on technology would be expensive, and limit our adaptability; we cannot produce sufficient tailor made technological solutions for all surgical eventualities we may encounter, and technology is intended to aid our craft and abilities, not replace it. Instead, we could improve our individual ability to lay secure, flat knots, in all circumstances and anatomical situations, and employ it universally. If a simple task as tying a secure knot can be so affected by technique, should we consider assessing the technique of all our manoeuvres and procedures?

## Article Summary

The majority of knots tied with a flat reef knot technique appeared to be secure.

All knots tied with the other two techniques, with all materials, slipped on testing and slipped markedly.

The length of suture material within loops held by flat reef knots was greater than in those held by knots tied with the other two techniques, suggesting these latter two slipped tighter during knot formation

Given the marked difference in the security of knots tied by the techniques studied, even though a relatively small number of knots were tied and assessed in this study, all clinicians and health professionals engaged in any invasive procedures should assess their knot tying techniques to ensure their knots are secure.

## Data Availability

All data available on request

## Author contributions

**Eric Drabble** Consultant surgeon and main author, performed procedures in study, data and statistical analysis, study design, reference collection, provision of figures and diagrams, photography of diagrams

**Sofia Spanopolou** Study design, performed procedures in study, vetting of paper, reference collection, provision of figures and diagrams, photography of diagrams

**Ellena Sioka** Facilitated study location and equipment, data collection and analysis, statistical analysis, vetting of paper, data collection

**Ellie Politaki** Performed procedures in study, vetting of paper, reference collection

**Isminis K Paraskeva** Performed procedures in study, vetting of paper, reference collection

**Effrosyni Palla** Performed procedures in study, vetting of paper, reference collection, provision of technical equipment

**Konstantinos G Railis** Technical support, data collection and storage, photography and videoing of procedures, analysis of photographic and video evidence

**Lauren Stockley** Statistical analysis and advice, provision of statistical packages and data plots

**Dimitris Zacharoulis** Professor of Surgery, University of Thessaly, academic lead for the project, responsible for providing location for study and technical support, support of design of study, vetting of paper.

## Acknowledgements

We are very grateful to the governing body and staff of IASO Thessaly, Larissa, Greece for providing a facility and technical support to allow this study to be performed, and for their encouragement of and enthusiasm for this project.

We wish to acknowledge Konstantinos G Railis for his technical support in filming and photographing during the course of this project, and for providing data storage. His support was instrumental in allowing us to achieve this study

We are also grateful to Ethicon Greece for providing the suture materials for this study

We are grateful to Kara Stevens of the University of Plymouth for her advice on our statistical analysis and presentation of our paper

We are grateful to, and acknowledge, the invaluable advice and historical knowledge of our surgical colleague, Sarandos Kaptanis, who guided us to appreciate the centuries old practice and wisdom of ancient Greek surgeons, who acknowledged and recorded the importance of secure knots, and sound technique two millenia before our study.

## Funding

‘This research received no specific grant from any funding agency in the public, commercial or not-for-profit sectors’.

**Email addresses:**

**Eric Drabble**

**Sofia Spanopolou**

**Ellena Sioka**

**Ellie Politaki**

**Isminis K Paraskeva**

**Effrosyni Palla**

**Konstantinos G Railis**

**Lauren Stockley**

**Dimitris Zacharoulis**

## Copyright

The Corresponding Author has the right to grant on behalf of all authors and does grant on behalf of all authors, an exclusive licence (or non exclusive for government employees) on a worldwide basis to the BMJ Publishing Group Ltd to permit this article (if accepted) to be published in BMJ editions and any other BMJPGL products and sublicences such use and exploit all subsidiary rights, as set out in our licence.

I affirm that this manuscript is an honest and transparent report of the study undertaken and of its results; no important aspecrt.

Eric Drabble – lead author

## References

1) F W Taylor Annals of Surgery: 1938: 107 (3): 458–468 Surgical knots

2) A M Gillen. A S Munsterman R R Hanson Veterinary Surgery. 2016. Vol. 45 issue 8. Pages 1034–1040 In vitro evaluation of the size, knot holding capacity and knot security of forwarder knot compared to square and surgeon’s knots using large gauge suture

3) A M Gillen. A S Munsterman. R Farag M O D Coleridge R R Hanso Veterinary Surgery. 2017. Vol. 46. Issue 2. Pages 297–305 In vitro evaluation of square and surgeon’s knots in large gauge sutures

4) R C Dinsmore J Am Coll Surg. 1995. Vol. 180. Issue 6. Pages 689–699 Understanding surgical knot security: a proposal to standardise the literature

5) E D Fong. A S Bartlett S Malak I A Anderson ANZ J. Surg. 2008. Vol. 78. Issue 3. Pages 164–166 Tensile strength of surgical knots in abdominal wound closure

6) S Jianmongkol G Hooper W Kowsumon T Thammaroj Hand Surg. 2006. Issue 11. Pages 93–99 A comparative bio mechanical study of the looped square slip knot and the simple surgical knot

7) J E Brouwers H Oosting D de Haas P J Klopper Surg Gynecol Obstet. 1991. Vol. 173. Issue 6. Pages 443–448 Dynamic loading of surgical knots

8) S S Kadirkamanathan J C Shelton C C Hepworth J G Laufer C P Swain J Am Coll Surg. 1996. Vol. 182. Issue 1. Pages 46–54 A comparison of the strength of knots tied by hand and at laparoscopy

9) H Tera. C Aberg Acta Chir Scand. 1976: 142 (1): 1–7 Tensile strengths of twelve types of knot employed in surgery, using different suture materials

10) J B Trimbos Obstet Gynecol. 1984: 64 (2): 274–280 Security of various knots commonly used in surgical practice

11) J B Trimbos. E J Van Rijssel. P J Klopper Obstet Gynecol. 1986: 68 (3): 425–430 Performance of sliding knots in monofilament and multifilament suture material

12) F Komatsu. R Mori. Y Uchio J Orthop Sci. 2006: 11 (1): 70–74 Optimum surgical suture material and methods to obtain high tensile strength at knots: problems of conventional knots and the reinforcement effect of adhesive agent

13) O Schadt M Glyde R E Day Veterinary Surgery 2010. Vol. 39. Issue 5. Pages 553–560 In vitro comparison of secure Aberdeen and square knots with plasma and fat coated polydioxanone

14) J D James. M M Wu. E K Batra. G T Rodeheaver. R F Edlich J Emerg Med: 1992: 10 (4): 469–480 Technical considerations in manual and instrument tying techniques

15) E Rosin. G M Robinson Vet Surg: 1989: 18 (4): 269–273 Knot Security of suture materials

16) A C Lee. R R Fahmy. G B Hanna World J Surg: 2008: 32 (12): 2736–2741 Objective evidence for optimum knot configuration

17) G Abbi. L Espinoza. T Odell. A Mahar. R Pedowitz Arthroscopy: 2006: 22 (1). 38–43 Evaluation of 5 knots and 2 suture materials for arthroscopic rotator cuff repair: very strong sutures can still slip

18) A D Shaw. G S Duthie J R Coll Surg Edinb: 1995: 40 (6): 388–391 A simple assessment of surgical sutures and knots

19) F J Van Rifssel. J B Trimbos. M H Booster Am J Obstet Gynecol: 1990: 162 (1): 93–97 Mechanical performance of square knots and sliding knots in surgery: comparative study

20) E Rosin. G M Robinson Vet Surg: 1989: 18: 269–273 Knot security of suture materials

21) P M Stoff. L G Ripley. M A Lavelle Ann R Coll Surg Engl: 2007: 89 (7): 713–717 The ultimate Aberdeen knot

22) J E Tidwell. V L Kish. J B Samora. J Prudhomme Orthopedics: 2012: 35 (4): e532–537 Knot security: how many throws does it really take?

23) X Avoine. B Lussier. V Brailovski. K Inaekyan. G Beauchamp Can J Vet Res: 2016: 80 (2): 162–170 Evaluation of the effect of 4 types of knots on the mechanical properties of 4 types of suture materials used in small animal practice

24) J D Sedluck. V M Williams. J DeSimone. D Page. B C Ghosh Surg Laparosc Endosc: 1996: 6 (2): 144–146 Laparoscopic knot security

25) H A Evin. T T Bilden. B C Noonan. A C Chong Iowa Orthop J: 2019: 131–140 A biomechanical comparison of varying base knot configurations with different overhand/underhand combinations of reversing half hitches on alternating posts after basic instructional training

26) C R Lutchman. L H Leung. R Moineddin. H F Chew Cornea: 2014: 33 (4): 414–418 Comparison of tensile strength of slip knots with that of 3-1-1 knots using 10-0 nylon sutures

27) O F Duenas-Garcia. G M Sullivan. K Leung. K L Billiar. M K Flynn Int Urogynecol J: 2018: 29 (7): 979–985 Knot integrity using different suture types and different knot-tying techniques for reconstructive pelvic floor procedures

28) I K Y Lo. S S Burkhart K C Chan. K Athanasiou Arthroscopy: 2004: 20 (5). 489–502 Arthroscopic knots: determining the optimal balance of loop security and knot security

29) S Kuptniratsaikul. P Weerawit. K Kongrukgreatiyos. T Promsang J Orthop Res: 2016: 34 (10): 1804–1807 Biomechanical comparison of four sliding knots and three high strength sutures; loop security is much different between each combination

30) D C Meyer. E Bachmann. A Laedermann. G Lajtai. T Jentzsch Orthop Traumatol Surg Res: 2018: 104 (8): 1277–1282 The best knot and suture configurations for high strength suture material. An in vitro biomechanical study

31) I K Y Lo. E Ochoa Jr. S S Burkhart Arthroscopy: 2010: 26 (9 Suppl): S120–126 A comparison of knot security and loop security in arthroscopic knots tied with newer high-strength suture materials

32) A Romeo. L F Fernandes. G V Cervantes. R Botchorishvili C Benedetto. L Adamyan. A Ussia. A Nattiez. W Kando. P R Koninckx J Minim Invasive Gynecol: 2020: 27 (6): 1395–1404 Which knots are recommended in laparoscopic surgery and how to avoid insecure knots

33) A M Schneider.R A Pedowitz. D A Evans Simul Healthc: 2019: 14 (1). 29–34 Validation of the FAST workstation as an objective evaluator of hand-tied surgical knots

34) E Silver. R Wu. J Grady. L Song J Oral Maxillofac Surg: 2016: 74 (7): 1304–1312 Knot security – how is it affected by suture technique, material, size and number of throws

35) N Van Leeuwen. J B Trimbos Gynecol Surg: 2012: 9 (4): 433–437 Strength of sliding knots in multifilament resorbable suture materials

36) R E Sanders. C M Kearney C T Buckley F Jenner P E Brama Veterinary Surgery. 2015. Vol. 44 issue 6. Pages 723–730 Knot security of 5 metric (USP 2) sutures; influence of knotting technique, suture materials and incubation time for 14 days and 28 days in phosphate buffered saline and inflamed equine peritoneal fluid

37) M M Good. L B Good. D D McIntire. S A Brown. C Y Wai J Surg Educ: 2013: 70 (1). 156–160 Surgical knot integrity: effect of suture type and caliber, and level of residency training

38) T E Ind. J C Shelton. J H Shepherd BJOG: 2001: 108 (10): 1013–1016 Influence of training on reliability of surgical knots

39) C A Zimmer. J G Thacker. D M Powell. K T Bellian. D G Becker. G T Rodeheaver. R F Edlich J Emerg Med: 1991: 9 (3): 107–113 Influence of knot configuration and tying technique on the mechanical performance of sutures

40) S S Ching. C W Mok. Y X Koh. S M Tan. Y K Tan J Surg Educ: 2013: 70 (1): 48–54 Assessment of surgical trainees’ quality of knot tying

41) E K Batra. D A Franz. M A Towler. G T Rodeheaver. J G Thacker. C A Zimmer. R F Edlich J Appl Biomater: 1993: 4(3): 241–247 Influence of surgeon’s tying technique on knot security

42) T M Muffly. C Cook. J Distasio. A J Bonham. R E Blandon J Surg Educ: 2009: 66 (5): 276–280 Suture end length as a function of knot integrity

43) D A Davis. D M Pellowski. E J Rawdon Dermatol Surg: 2013: 39 (5): 729–733 All monofilament sutures assume sliding confirmation in vivo

44) C C Annunziata. D B Drake. J A Woods A J Gear. G T Rodeheaver. R F Edlich J Emerg Med: 1997; 15 (3): 351–356 Technical considerations in knot construction. Part 1 continuous percutaneous and dermal suture closure

45) J H Maddocks J B Keller Ropes in equilibrium Journal of Applied Mathematics. 1987. Vol. 47. Pages 1185–1200

46) Crowell The physics of knots Webpage http://www.lightandmatter.com/article/knots.html

47) J J Ivy. J B Unger J Hurt. D Mukherjee Am J Obstet Gynecol: 2004: 191 (5): 1618–1620 The effect of number of throws on knot security with non-identical sliding knots

48) J J Ivy. J B Unger. J Hurt. D Mukherjee Am J Obstet Gynecol: 2004: 191 (5): 1618–1620 Th effect of number of throws on knot security with non-identical sliding knots

49) J J Ivy. J B Unger. D Mukherjee Am J Obstet Gynecol: 2004: 190 (1): 83–86 Knot integrity with non-identical and parallel sliding knots

50) J J Trimbos. E J Vankijssel. P J Klopper Obstet Gynecol: 1986: 68 (3): 425–430 Performance of sliding knots in monofilament and multifilament suture material

51) T M Muffly N Kow I Iqbal M D Barber J. Surg Education 2011. Vol. 68. Issue 2. Pages 130–133 Minimum number of throws needed for knot security

52) Participants’ handbook of the sixth intercollegiate Basic Surgical Skills course RCS Eng Education Department 2017

53) D A Franz. E K Batra. R F Morgan. R F Edlich Orthopaedics: 1995: 18 (6): 555–558 A portable tensiometer for assessment knot tying technique

54) B T Hanypsiak. J M DeLong. L Simmons. W Lowe. S Burkhart Am J Sports Med: 2014: 42 (8): 1978–1984 Knot strength varies widely among expert arthroscopists

55) A F Burton. C Horstman. D R Mason Vet Comp Orthop Traumatol: 2015: 28 (6): 391–400 Tensions generated in lateral fabellotibial suture model. Comparison of methods of application of tension, fixation of tension and suture material

56) T M Muffly. LM Espaillat-Rijo. AM Edwards. A Horto J Surg Educ: 2012: 69 (2): 215–217 Operating room fatigue: is your twentieth surgical knot as strong as your first?

57) I K Stone. J A Von Fraunhofer. B J Masterson Am J Obstet Gynecol: 1985: 151 (8): 1087–1093 A comparative study of suture materials: chromic catgut and chromic catgut treated with glycerin

58) Z Babetty. A Suemer S Altintas. S Ergueney. S Goeksel Arch Surg: 1998: 133 (7). 727–734 Changes in knot holding capacity of sliding knots in vivo and tissue reaction

59) J G Thacker G Rodeheaver. L Kurtz. MT Edgerton. RF Edich Am J Surg 1977: 133 (6): 713–715 Mechanical performance of sutures in surgery

60) L Fischer. T Bruckner. PB Mueller-Stitch. J Hoer. HP Knaebel. MW Buchler. CM Saler Langenbecks Arch Surg: 2010: 395 (4): 445–450 Variability of surgical knot tying techniques: do we need to standardise?

61) S S Ching. CW Mok. YX Koh. S-M Tan. Y K Tan J Surg Educ: 2013: 70 (1): 48–54 Assessment of surgical trainees’ quality of knot tying

62) S H Kim. D Glaser. J Doan. SW Chung. HY Choi. JH Oh. AR Hargens Arthroscopy: 2013: 29 (8): 1380–1386 Loop securities of arthroscopic sliding knot techniques when the suture loop is not evenly tensioned

63) J B Trimbos. A Niggebrugge. R Trimbos. E J Van Rijssel Eur J Surg: 1995: 161 (5): 319–322 Knotting abilities of a new absorbable monofilament suture: poliglecaprone 25 (monocryl)

64) H Li W Lei J Gastroint Surg 2019: 23 (3): 617 Laparoscopic extended left hemi-hepatectomy plus caudate lobectomy for caudate lobe hepatocellular carcinoma

65) S J Tharakan. D Hiller. RM Shapiro. SK Bose. T A Blinma Surg Endosc 2016: 30 (10): 4653 – 4658 Vessel sealing comparison: old school is still hip

66) C L Nota. IQ Molenaar. IH Borel Rinkes. J Hagendoor Surg Oncol 2020: 34: 206–207 Robotic liver resection of segment 7: A step-by-step description of the technique

67) J J Hage World Journal of surgery. 2008 32 (4) Pages 648–655 Heraklas on knots; sixteen surgical nooses and knots from the first century AD

68) F Palazzo Fundamentals of General Surgery 2018 Page 50 “Knot tying, Ligatures, and suturing”

69) J H Przytycki in “Knots, low-dimensional topology and applications” Knots in Hellas, International Olympic Academy Greece July 2016: Pages 115–119

70) D Steel “The elements and practice of rigging and seamanship” London 1794. Page 183

71) Lever. Darcy The Young Sea Officer’s Sheet Anchor (2nd edition) 1998 1819 Mineola. NY: Dover Publications Page 83 ISBN 978-0-486-40220-8

72) R H Dana “The Seaman’s Friend: A Treatise on Practical Seamanship” (14th revised and corrected ed.) 1997 1879 Mineola NY: Dover Page 49 ISBN 0-486-29918-X

73) J R Miller. CR Deeken. S Ray. MC Henn. TS Lancaster. RB Schuessler. RJ Danniano. SJ Melby Ann Thorac Surg: 2015: 100 (6) 2325–2329 Expanded polytetrafluoroethylene for chordal replacement: preventing knot failure

74) A D Shaw. GS Duthie J R Coll Surg Edinb: 1995: 40 (6). 388–391 A simple assessment of surgical sutures and knots

75) P Petit “Why knot?: How to tie more than sixty ingenious, useful, beautiful, lifesaving, and secure knots! 2013. Pages 80–83 “The Square Knot (revisited)”

76) B S Gupta. KW Wolf. RW Postlethwait Surg Gynecol Obstet: 1985: 161 (1): 12–26 Effect of suture material and construction on frictional properties of sutures

